# Metropolitan Wastewater Analysis for COVID-19 Epidemiological Surveillance

**DOI:** 10.1101/2020.04.23.20076679

**Authors:** Walter Randazzo, Enric Cuevas-Ferrando, Rafael Sanjuán, Pilar Domingo-Calap, Gloria Sánchez

**Author notes:** corresponding authors Correspondence to: Pilar Domingo-Calap; Gloria Sánchez.

## Abstract

**Background:** The COVID-19 disease, caused by severe acute respiratory syndrome coronavirus 2 (SARS-CoV-2), is a rapidly emerging pandemic which has enforced extreme containment measures worldwide. In the absence of a vaccine or efficient treatment, cost-effective epidemiological surveillance strategies are urgently needed.

**Methods:** Here, we have used RT-qPCR for SARS-CoV-2 detection in a series of longitudinal metropolitan wastewaters samples collected during the earliest stages of the epidemic in the Region of Valencia, Spain.

**Results:** We were able to consistently detect SARS-CoV-2 RNA in samples taken when communicated cases in that region were only incipient. We also find that the wastewater viral RNA context increased rapidly and anticipated the subsequent ascent in the number of declared cases.

**Interpretation:** Our results strongly suggest that the virus was undergoing community transmission earlier than previously believed, and show that wastewater analysis is a sensitive and cost-effective strategy for COVID-19 epidemiological surveillance. Routine implementation of this surveillance tool would significantly improve our preparedness against new or re-occurring viral outbreaks.

---

The emergence of severe acute respiratory syndrome coronavirus 2 (SARS-CoV-2) in Wuhan, China, has rapidly led to a pandemic scenario, with over 2·6 million COVID-19 confirmed cases globally as of April 23, 2020. COVID-19 symptoms are varied and often non-specific, including fever, cough, and diarrhea, among others. A non-negligible percentage of infected people develop pneumonia, which can subsequently lead to severe respiratory distress requiring mechanical ventilation, organ failure, viral sepsis, and death^1^. The widespread nature of the pandemics and the lack of easy symptom-based diagnosis, treatment, or vaccine has enforced drastic and extremely costly epidemiological control measures including worldwide lockdowns. Whereas massive RT-qPCR testing campaigns are being deployed in many countries to assess the actual prevalence of the virus, this is not a feasible surveillance strategy for the general population over the long term.

Spain now stands as the second most extensively affected country worldwide with over 200,000 confirmed COVID-19 infections and more than 20,000 deaths. The first three confirmed cases in the Iberian Peninsula were communicated on February 25, 2020 in Madrid, Barcelona, and Villareal, a small town nearby the city of Valencia. Furthermore, a retrospective analysis carried out in March showed that the first death in Spain from COVID-19 actually occurred on February 13 in Valencia. Therefore, the Valencian Region constituted the earliest known COVID-19 spot in Spain^2^. However, at the time, it was assumed that no community transmission was ongoing and, as a result, major containment measures were not enforced until March 15.

Although SARS-CoV-2 is primarily a respiratory, airborne virus, previous studies with the related SARS-CoV-1 (the causative agent of the 2003 SARS outbreak) suggested the possibility of fecal-oral transmission based on detection of viral RNA by RT-qPCR in the stools of patients^3^. Recent studies indicate that SARS-CoV-2 can be also excreted in feces in asymptomatic carriers and in recently recovered patients^4–10^. Specifically, viral RNA was detected in feces up to 10 days after viral clearance from the respiratory tract, regardless of disease severity^11^. This implies that wastewaters may contain viral particles or viral RNA that could be used as an epidemiological surveillance tool. Wastewaters can also collect viruses present in the oral cavity and upper respiratory tract that are shed during personal hygiene. Compared to systematic testing of individuals, wastewater analysis is obviously less invasive, simpler and cheaper, but the sensitivity and reliability of this method remains to be shown. Previous work has established similar methods for the epidemiological surveillance of enteric viruses including norovirus, hepatitis A virus, influenza, and poliovirus^12–14^, and recent opinion letters^15-17^ and ongoing work^18–21^ suggest that COVID-19 detection in sewage is technically feasible, based on preliminary results obtained from a limited number of samples in China, Australia, the Netherlands, and USA.

Here, we show that SARS-CoV-2 can be reproducibly detected by RT-qPCR in longitudinal samples from sewage treatment plants that receive wastewaters from over one million inhabitants in the metropolitan area of Valencia, Spain. We analyzed 15 samples taken between February 12 and April 14, 2020 at three wastewater treatment plants. Following concentration of viral content by flocculation, a standard RT-qPCR procedure allowed us to detect SARS-CoV-2 RNA in 12/12 samples collected from March 9 to April 14, 2020, with Ct values ranging between 34·00 and 37·84, correspondingly revealing between 5·22 and 5·99 log_10_ genomic copies (gc)/L (Table 1). SARS-CoV-2 RNA was not detected in a single sample from February 12, but was detected in one of the two samples collected in February 24 for RT-qPCR region N2, whereas region N1 remained negative for this time point. Hence, samples from February 24 provide the earliest piece of evidence that the virus was circulating in the community. Interestingly, we consistently detected SARS-CoV-2 RNA in samples collected on March 9 and March 11, when only 50 and 76 cumulative cases were declared in the entire Region of Valencia. This validates wastewater RT-qPCR analysis as a sensitive and reliable technique for early detection of SARS-CoV-2 outbreaks.

**Table 1.** Detection of SARS-CoV-2 RNA by RT-qPCR in wastewater samples from three treatment plants in the metropolitan region of Valencia, Spain.

| Sampling date (2020) | Water treatment plant | Ct ± SD (N1)<sup>a</sup> | Log<sub>10</sub> gc/L ± SD (N1)<sup>a</sup> | Ct ± SD (N2)<sup>b</sup> | Log<sub>10</sub> gc /L ± SD (N2)<sup>b</sup> |
| --- | --- | --- | --- | --- | --- |
| February 12 | Quart-Benàger | nd<sup>c</sup> | nd<sup>c</sup> | nd<sup>c</sup> | nd<sup>c</sup> |
| February 24 | Pinedo 1 | nd<sup>c</sup> | nd<sup>c</sup> | nd<sup>c</sup> | nd<sup>c</sup> |
|  | Pinedo 2 | nd<sup>c</sup> | nd<sup>c</sup> | 37.84<sup>d</sup> | 5.22<sup>d</sup> |
| March 9 | Pinedo 1 | 35.16<sup>d</sup> | 5.4<sup>d</sup> | 36.88<sup>d</sup> | 5.45<sup>d</sup> |
| | Pinedo 2 | 35.28 ± 0.87 | 5.37 ± 0.26 | 35.55<sup>d</sup> | 5.77<sup>d</sup> |
| March 11 | Quart-Benàger | 34.64 ± 0.23 | 5.56 ± 0.07 | 36.95 ± 0.00 | 5.43 ± 0.00 |
| April 6 | Pinedo 1 | 34.75 ± 0.09 | 5.53 ± 0.03 | 34.70 ± 0.01 | 5.98 ± 0.00 |
| | Pinedo 2 | 34.67 ± 0.20 | 5.55 ± 0.06 | 36.48 ± 0.66 | 5.55 ± 0.16 |
| | Quart-Benàger | 34.53 ± 0.93 | 5.59 ± 0.28 | 34.65 ± 0.67 | 5.99 ± 0.16 |
| April 9 | Pinedo 1 | 35.38 ± 0.91 | 5.34 ± 0.27 | 36.11 ± 0.78 | 5.64 ± 0.19 |
| | Pinedo 2 | 34.93 ± 0.78 | 5.47 ± 0.23 | 35.71 ± 0.28 | 5.73 ± 0.07 |
| April 11 | Quart-Benàger | 34.45 ± 0.19 | 5.62 ± 0.06 | 35.11 ± 0.59 | 5.88 ± 0.14 |
| April 13 | Quart-Benàger | 34.00 ± 0.11 | 5.75 ± 0.03 | 34.89 ± 0.33 | 5.93 ± 0.08 |
| April 14 | Pinedo 1 | 35.10 ± 0.50 | 5.41 ± 0.15 | 35.82<sup>d</sup> | 5.71<sup>d</sup> |
| | Pinedo 2 | 35.48 ± 0.40 | 5.31 ± 0.12 | 35.61 ± 0.69 | 5.76 ± 0.17 |

<sup>a</sup>PCR region encompassing 2019-nCoV\_N1 Combined Primer/Probe Mix.
<sup>b</sup>PCR region encompassing 2019-nCoV\_N2 Combined Primer/Probe Mix.
<sup>c</sup>Not detected (Ct > 40).
<sup>d</sup>Based on a single technical replicate.

For each of the samples collected in April, we also analyzed treated wastewaters, which are routinely discharged to the sea or used for irrigation purposes. We found no evidence of viral RNA in 9/9 samples. These samples provide an additional negative control for our RT-qPCR analysis, reinforcing the conclusion that the signal obtained in untreated waters unlikely corresponds to background signal or non-specific DNA amplification. Importantly, these results confirm that current wastewater treatment procedures efficiently clear the virus. Worryingly, waterborne transmission may occur in developing countries where sewage treatment is far less efficient, although no evidence of fecal-oral transmission has been validated at the moment.

In conclusion, our results show that the virus was probably undergoing local community transmission by the time the very first cases were declared in the Region of Valencia (Figure 1). This contrasts with the previously accepted view that, in late February and early March, essentially all COVID-19 cases in Spain were imported or directly traceable through contacts, and that there was no ongoing community transmission at the time. We also find that the RT-qPCR signal in wastewaters increased and reached a plateau faster than declared cases. These results strongly suggest that analysis of wastewaters by RT-qPCR analysis is an efficient strategy for the epidemiological surveillance of COVID-19.

**Figure 1.**
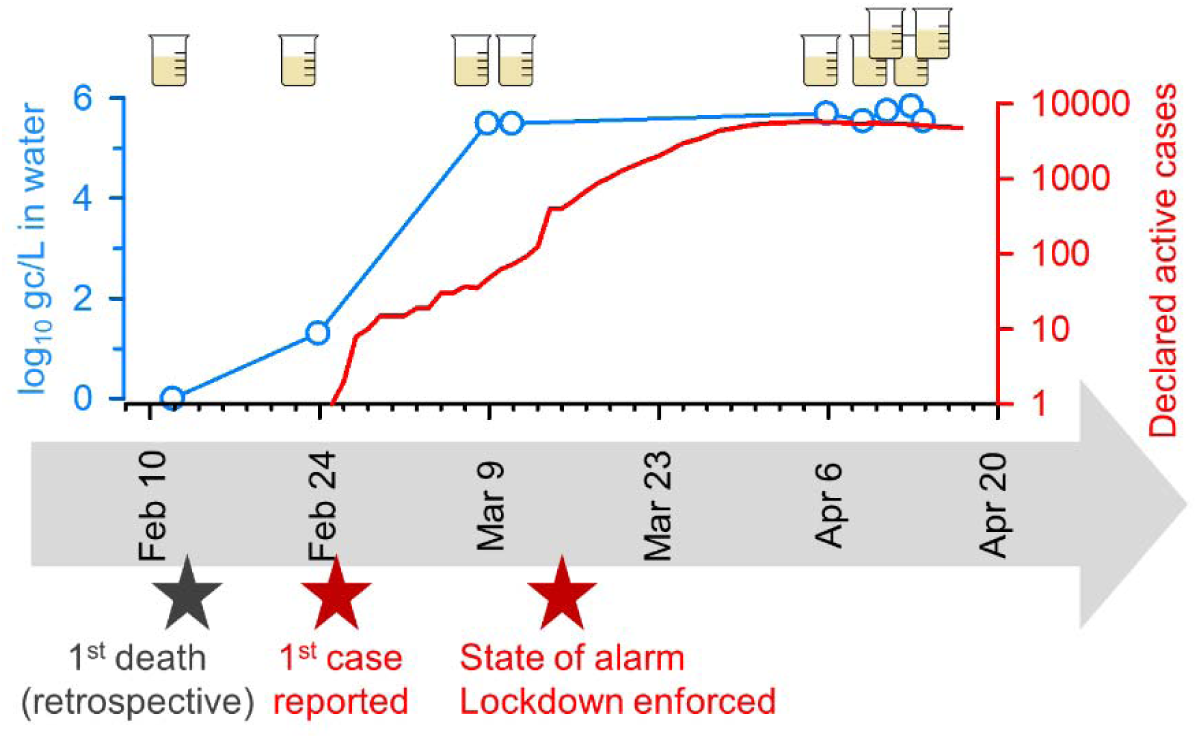
Series of events and SARS-CoV-2 epidemiological follow-up by wastewater RT-qPCR. Blue: Evolution of mean viral load values as inferred by RT-qPCR. For each sampling time point (indicated with beakers), log viral load data shown on Table 1 were averaged across treatment plants and PCR regions. Samples with undetectable RNA levels were arbitrarily assigned a log viral load of zero for averaging purposes. Red: Evolution of the number of active declared cases in the Region of Valencia. Bottom: series of events related to the COVID-19 pandemics in the Valencian Region.

Extreme lockdown measures are currently allowing Spain and other countries to partially mitigate SARS-CoV-2 spread and may help us reduce disease prevalence within the next weeks or months. However, inevitable relaxation of current containment measures may lead to recurring local outbreaks or case imports from other regions. As such, it is of extreme importance to set up feasible and reliable epidemiological surveillance strategies that will improve our preparedness in the event of future viral re-emergencies.

## Methods

### Wastewater sampling

Samples of metropolitan wastewater from Valencia (Spain) were taken at different time points in a two-month longitudinal study spanning from February 12 to April 14, 2020. Specifically, samples were taken from wastewater treatment plants Pinedo 1, Pinedo 2 and Quart-Benàger, all belonging to the Empresa Pública de Saneamiento de Aguas Residuales (Generalitat Valenciana). Some samples were collected before and after wastewater treatment, and all samples were kept at 4°C until analysis. Previous studies have shown that SARS-CoV-2 is stable at 4°C for at least 14 days^19,22^. Since our earliest samples remained stored at 4°C for almost two months, we cannot discard viral degradation. However, this would lead to underestimation of the viral RNA present in these samples, which ultimately would strengthen our conclusion that RT-qPCR wastewater analysis allows early detection of disease outbreaks.

### Sample processing and RNA extraction

Viral concentration was carried out by aluminum-driven flocculation. For this, 200 mL water samples were adjusted to pH 6·0 and an Al(OH)3 precipitate was formed by adding 1:100 v:v of 0·9 N AlCl3 solution. After pH readjustment to 6·0, samples were agitated slowly for 15 min at room temperature. Precipitates were collected by centrifugation at 1,700 × g for 20 min. Pellets were resuspended into 10 mL of 3% beef extract (pH 7·4), and samples were shacked for 10 min at 150 rpm^23^. A concentrate was then formed by centrifugation at 1,900 × g for 30 min and the pellet resuspended in 1 mL of PBS^24^. As a process control, samples were spiked with 10^5^ PCR units of mengovirus vMC0 (CECT 100000) according to ISO 15216-2:2017. RNA extraction was performed using the Nucleospin RNA virus Kit (Macherey-Nagel) following the recommended protocols and using a Plant RNA Isolation Aid (Ambion) pre-treatment^25^.

### SARS-CoV-2 RT-qPCR

The presence of SARS-CoV-2 was determined using the PrimeScript™ One Step RT-PCR Kit and the RT-qPCR diagnostic panel assays validated by the US Centers for Disease Control and Prevention (2019-nCoV RUO Kit) using the positive control (2019-nCoV_N_Positive Control) provided by IDT (Integrated DNA Technologies). The RT-qPCR was carried out following the manufacturer’s instructions, recommended standards, and positive controls in a LightCycler 480 (Roche Diagnostics) instrument. Each RNA extract was analyzed in duplicate. Undiluted and tenfold diluted RNA extracts were analyzed to account for the presence of RT-qPCRs inhibitors. A calibration curve was performed using the 2019-nCoV_N_Positive Control provided by IDT. For each RT-qPCR run, a series of three positive and negative controls (extraction and PCR) were included. Cycle threshold (Ct) values were used to calculate gc/L in the original sample. Ct values lower than 40 were considered positive for SARS-CoV-2, as proposed previously^9^. In all cases, the internal mengovirus control showed recovery rates >1%, which conformed to ISO15216-2:2017 standards.

### Epidemiological data

The sewage treatment plants under study collected wastewaters from approximately 1,200,000 inhabitants in 22 townships of the Valencian metropolitan area (Alaquas, Albal, Alcasser, Aldaia, Alfafar, Benetusser, Beniparell, Burjassot, Catarroja, Llocnou de la Corona, Manises, Massanassa, Mislata, Paiporta, Picanya, Picassent, Quart de Poblet, Sedavi, Silla, Torrent, Valencia, and Xirivella). The number of declared active cases per day for the Region of Valencia was obtained from Conselleria de Sanidad Universal y Salud Pública (Generalitat Valenciana).

## Data Availability

All data are available in the manuscript.

## Acknowledgments

We thank the Generalitat Valenciana and the Empresa Pública de Saneamiento de Aguas Residuales (EPSAR) for providing access to wastewater. This research was funded by CSIC 202070E101 grant and AGL2017-82909 (AEI/FEDER, UE) funded by Spanish Ministry of Science, Innovation and Universities to G.S. and ERC Consolidator Grant 724519-Vis-à-Vis to R.S. W.R. was supported by APOSTD/2018/150 postdoctoral fellowship, and E.C-F. was supported by a predoctoral contract from the MICINN, Call 2018.

## Author contributions

W.R. performed the experiments and analyzed data; E.C-F. performed the experiments; R.S. analyzed data and co-wrote manuscript; P.D-C. conceived study, obtained the samples, and co-wrote manuscript; G.S. conceived study and supervised work. All authors have read and agreed to the published version of the manuscript.

